# Instantaneous Wave-Free Ratio—Guided vs Angiography-Guided Coronary Artery Bypass Grafting: 36-Months Graft Patency and Clinical Outcomes of a Randomized Trial

**DOI:** 10.64898/2026.04.01.26350013

**Authors:** Rasa Ordiene, Ramunas Unikas, Rimantas Benetis, Povilas Jakuska, Indre Ceponiene, Antanas Jankauskas, Ali Aldujeli, Jurgita Plisiene, Aiste Ivanauskiene, Tautvydas Kabosis, Zilvinas Krivickas, Prakash P Punjabi, Justin Davies

## Abstract

**Background:** Coronary artery bypass grafting (CABG) to physiologically non-significant coronary artery stenosis may result in graft failure due to competitive native flow. We evaluated whether an instantaneous wave-free ratio (iFR)-guided revascularization strategy improves graft patency and clinical outcomes compared to conventional angiography-guided CABG.

**Methods:** In this prospective, randomized, single-blinded trial, patients with multivessel disease and at least one angiographically intermediate stenosis (50%–75%) were randomized to either CABG guided by angiography alone or angiography supplemented with iFR assessment groups. The primary endpoint was graft patency (occlusion or hypoperfusion of the graft) evaluated by coronary computed tomography angiography (CCTA) at 2, 12, and 36 months.

**Results:** At 36 months, 78% of the patients completed follow-up. Left internal mammary artery (LIMA)-to-left anterior descending (LAD) artery graft patency was significantly higher in the iFR-guided group than in the angiography-guided group (80.5% vs. 56.8%; absolute risk difference, 23.7% [95% *CI*, 3.7%–43.8%]; *RR*, 1.42 [95% *CI*, 1.03–1.95]; *P* = 0.03). Saphenous vein graft patency also improved with iFR guidance (90.2% vs. 70.3%; *P* = 0.046). Major adverse cardiac and cerebrovascular events (MACCE) were similar between groups (28% vs. 20%; *RR*, 1.40 [95% *CI*, 0.69–2.85]; *P* = 0.48).

**Conclusions:** iFR-guided CABG advocates significantly improved mid-term graft patency compared with angiography-guided CABG by optimizing surgical target selection and reducing competitive flow.

**CLINICAL PERSPECTIVE:** *What Is New?:*

- This prospective randomized trial demonstrates that physiological guidance using instantaneous wave-free ratio (iFR) significantly improves both arterial and venous graft patency at 36 months follow up compared to angiography-guided coronary artery bypass grafting (CABG).
- A pre-operative iFR threshold of >0.875 in the target vessel was identified as a potent predictor of subsequent graft occlusion or hypoperfusion.
- The study provides longitudinal evidence that competitive flow, identified by iFR, remains a critical determinant of long-term conduit durability.

*What Are the Clinical Implications?:*

- Integrating routine iFR assessment into the surgical planning for multivessel coronary artery disease may identify fewer significant stenoses, reducing the need for CABG.
- Using physiological rather than purely anatomical criteria for revascularization may mitigate the risk of graft failure associated with competitive native flow.
- These results suggest that preoperative physiological assessment can refine surgical revascularization strategies and enhance long-term conduit durability.

## INTRODUCTION

Approximately 400,000 coronary artery bypass graft (CABG) surgeries are being performed annually, making it one of the most common surgical procedures worldwide (1). According to the 2021 ACCF/AHA/AATS/STS/SCAI guidelines, CABG remains the gold standard for revascularization—offering superior survival rates and a reduction in myocardial infarction—compared with percutaneous coronary intervention (PCI) in patients with complex or diffuse multivessel coronary artery disease (CAD) (2,3). However, graft failure remains a significant limitation, impacting long-term clinical outcomes and necessitating repeat revascularization. The most frequently used conduits, saphenous vein grafts (SVG), fail in 2%–25% of cases at 1 year and 40%–50% at 10 years or more (4). Other studies indicate overall graft failure rates of 5% within one week, 16% at 1 year, 20% at 3 years, 31% at 6 years, and 49% at 10 years (5). Furthermore, studies based on fractional flow reserve (FFR) demonstrated that in functionally significant lesions, 13.7% of arterial and 5.9% of venous conduits were occluded after 1 year; conversely, in functionally non-significant lesions, occlusion rates were 21.9% and 20.0%, respectively, meanwhile patency rates for left internal mammary artery (LIMA) grafts were 93.8%, while radial artery conduits achieved 71% (6).

Beyond technical factors, a primary driver of graft failure is the progression of atherosclerosis and intimal hyperplasia (IH) (7). A key mechanism contributing to this process is competitive flow between the native coronary circulation and the bypass graft, particularly when grafts are placed on physiologically non-significant coronary stenoses (6,8). Because angiography alone may overestimate lesion severity, it often results in the grafting of lesions that do not significantly impair coronary flow. To avoid the placement of unnecessary grafts, it is hypothesized that functional assessment of coronary artery stenosis prior to CABG could be beneficial (9).

To assess the hemodynamic relevance of intermediate-grade stenosis (typically 50%–75%), fractional flow reserve (FFR) or instantaneous wave-free ratio (iFR) are recommended (Class IA recommendation, ESC/EACTS Guidelines on Myocardial Revascularization) (10). Physiological assessment using iFR allows to evaluate the stenoses without pharmacological hyperaemia and has demonstrated clinical utility in guiding PCI. However, its role in guiding surgical revascularization remains incompletely defined (11). Various studies suggest diverse results; non-randomized single-center registry studies show that hemodynamically insignificant stenoses predispose to graft failure (12). Furthermore, studies based on preoperative FFR evaluation showed a significantly higher rate of patent arterial grafts at 6-year follow-up if the target coronary artery had a hemodynamically significant lesion (6). However, the GRAFFITI trial failed to show any difference in graft patency between FFR-guided and angiography-guided CABG groups (13). A meta-analysis by Bruno et al. revealed that FFR-guided surgical revascularization is associated with fewer grafts and shorter procedural times, but no difference in major adverse cardiac and cerebrovascular events (MACCE) (14). Retrospective research in Japan determined that even when the FFR is positive (<0.80), graft failure is significantly more frequent when conduits are anastomosed to vessels with physiologically non-significant iFR (>0.89) (15).

Consequently, there is a gap in evidence regarding whether iFR-guided CABG is beneficial in terms of graft patency and clinical outcomes. This pilot, single-center, blinded study was designed to prospectively compare graft patency and clinical outcomes using a physiologically guided approach versus standard angiographically guided CABG at 36-month follow-up. We hypothesized that iFR-guided CABG would improve graft patency by identifying physiologically significant lesions and avoiding the grafting of non-flow-limiting stenoses.

## METHODS

### Study Design and Population

This was a prospective, randomized, single-blind, single-center controlled trial conducted at the Hospital of the Lithuanian University of Health Sciences Kaunas Clinics between 2018 and 2021. The study compared an angiography-guided coronary artery bypass grafting (CABG) strategy with an instantaneous wave-free ratio (iFR)-guided approach. Patients aged 18 years or older with stable angina and multivessel coronary artery disease (CAD), defined as significant disease in two or more major epicardial coronary arteries or their branches, were eligible. Enrolment required the presence of at least one intermediate coronary stenosis (50%–75% by visual or quantitative assessment).

The Heart Team recommended CABG as the optimal revascularization strategy for all participants. Of 350 patients evaluated, 110 met the eligibility criteria. Strict inclusion and exclusion criteria were applied to minimize confounding effects on major adverse cardiac and cerebrovascular events (MACCE) (Table 1). The study was approved by the Kaunas Regional Biomedical Research Ethics Committee (BE-2-89), and all participants provided informed consent.

**Table 1.** Detailed Inclusion and Exclusion Criteria for Participant Selection in the iCABG Study.

| Inclusion criteria | Exclusion criteria |
| --- | --- |
| 1. Age over 18 years old.<br>2. Willing to participate and able to understand, read and sign the informed consent document before the planned procedure.<br>3. Eligible for coronary angiography and coronary artery bypass grafting.<br>4. Coronary artery disease in two or more native major epicardial vessels or their branches including LAD, with visually at least one artery with intermediate stenosis.<br>5. Stable angina.<br>6. Plan for CABG as optimal revascularization mode as decided by the treating clinical team. | 1. LV ejection fraction $\leq 40\%$.<br>2. History of atrial fibrillation / flutter.<br>3. History of MI within the last 30 days.<br>4. Emergency CABG.<br>5. Severe valvular heart disease.<br>6. Haemodynamic instability at the time of angiography.<br>7. Contraindications to CABG, had previously CABG.<br>8. Significant hepatic or lung disease (chronic pulmonary obstructive disease), and / or malignant disease with unfavorable prognosis that may influence survival within the next 5 years, history of chest radiotherapy.<br>9. Pregnancy.<br>10. STEMI within 48 hours of procedure.<br>11. Allergy to contrast media and renal failure. |

Abbreviations: CABG, coronary artery bypass grafting; LAD, left anterior descending artery; LV, left ventricular; MI, myocardial infarction; STEMI, ST-segment elevation myocardial infarction.

### Data Collection

Comprehensive data on patient demographics, comorbidities, and medication regimens were collected at baseline. Procedural data included coronary angiography findings and transthoracic echocardiography (TTE) results (ejection fraction, wall motion abnormalities, and valvular function). Clinical outcomes were categorized into short-term complications and long-term events, including ischemic or haemorrhagic stroke, myocardial infarction (MI), target vessel revascularization (TVR), cardiovascular death, and all-cause mortality. Data were extracted from inpatient and outpatient records and verified by two specialists.

### Physiological Assessment and Randomization

Coronary artery stenosis severity was evaluated using quantitative coronary angiography (QCA) and independently reviewed by two interventional cardiologists. Stenoses of 50%–75% were classified as intermediate.

Physiological assessment was performed using a pressure wire under resting conditions to determine iFR. After iFR measurement, patients were randomized (1:1) to either angiography-guided or iFR-guided CABG, irrespective of the iFR values. In the iFR-guided group, lesions with an iFR ≤0.89 were considered physiologically significant and targeted for grafting, while those with an iFR ≥0.90 were deferred.

Nine patients were excluded in post-iFR assessment due to ethical considerations (single-vessel significant disease identified), leading to a decision for percutaneous coronary intervention (PCI). The final 101 patients underwent the assigned surgical strategy. iFR measurements were blinded to the surgical team.

### Surgical Procedure

All patients underwent standard CABG using arterial and venous conduits according to institutional practice. A protocol-mandated exception was made for the left internal mammary artery (LIMA) to left anterior descending (LAD) artery graft, which was performed in all patients regardless of iFR values due to its established survival benefit. In the angiography-guided group, the number of grafts was determined by the surgeon based on QCA. In the iFR-guided group, grafting of non-LAD vessels was performed only for stenoses with an iFR ≤0.89.

### CT Imaging Protocol and Analysis

Coronary computed tomography angiography (CCTA) was performed at 2, 12, and 36 months post-CABG using a 320-detector (Aquilion One, Toshiba Medical Systems) or 256-detector scanner (Revolution, GE Healthcare) with prospective ECG gating. Imaging parameters included tube voltage of 100–120 kV and collimation of 0.6–0.625 mm, adhering to established guidelines (11). Axial images and multiplanar reconstructions were independently evaluated by a radiologist (12 years of experience) and a cardiologist (6 years of experience). Interpretation followed the Society of Cardiovascular Computed Tomography guidelines. Graft occlusion was defined as complete discontinuation of contrast flow; hypoperfusion or stenosis was identified by locally reduced contrast intensity or disrupted flow.

### Study Endpoints

The **primary endpoint** was the rate of graft occlusion or hypoperfusion at 2, 12, and 36-months post-CABG, characterizing the timing of graft failure. **Secondary endpoints** included the composite rate of MACCE (cardiovascular death, non-fatal MI, stroke, or TVR) and individual graft patency at 2, 12, and 36 months.

### Statistical Analysis

Statistical analysis was performed using SPSS version 27.0 (IBM Corp., Armonk, NY. The distribution of numerical variables was assessed using the Kolmogorov-Smirnov test. Parametric data are expressed as mean ± SD, while non-parametric data are presented as median and interquartile range (IQR, 25th–75th percentiles). Categorical variables were compared using the chi-square (χ^2^) or Fisher exact test, as appropriate. For continuous variables, differences between groups were assessed using the Student *t* test for normally distributed data or the Mann-Whitney *U* test for non-normally distributed variables. Multiple group comparisons of non-parametric data were performed using the Kruskal-Wallis test. Relative risks (RR) and 95% confidence intervals (CIs) were calculated to evaluate the strength of associations. Receiver operating characteristic (ROC) curves were generated to determine the diagnostic probability and optimal threshold of iFR values for predicting graft patency. The association between iFR and graft patency was further evaluated using binary logistic regression. Multivariable logistic regression analysis was performed to identify independent predictors of graft patency, adjusting for relevant clinical variables. Long-term clinical outcomes were analyzed using the Kaplan-Meier method, with differences between groups assessed via the log-rank test. Statistical significance was defined as a two-sided P < 0.05 for all analyses.

## Results

### Study Population and Procedural Characteristics

From December 2018 to June 2021, 110 patients were enrolled and randomly assigned to either the iFR-guided CABG group (n = 59) or the angiography-guided CABG group (n = 51). In the iFR-guided group, 26 patients (44.1%) were diagnosed with single- or double-vessel disease rather than multivessel disease based on physiological assessment. After unblinding iFR results in the iFR-guided arm, 9 of 59 patients (15.3%) did not proceed with CABG due to physician or patient preference, while 17 patients (28.8%) chose to undergo CABG despite the findings. One patient in the angiography-guided group died in the immediate postoperative period due to acute mesenteric thrombosis. Consequently, the final analysis included 100 patients who underwent surgical revascularization. Comprehensive demographic and clinical characteristics are detailed in Table 2.

**Table 2.** Clinical Characteristics of the Patients.

|  | Angiography guided<br>CABG group (*n*=50) | iFR<br>guided CABG<br>group (*n*=50) | *P* value |
| --- | --- | --- | --- |
| Male gender, *n* (%) | 37 (74%) | 40 (80%) | 0,476 |
| Age, years (*SD*) | 65,7(8,3) | 65,1(8,3) | 0,71 |
| Hypertension, *n* (%) | 49 (98%) | 49 (98%) | 1,0 |
| Diabetes mellitus, *n* (%) | 13 (26%) | 9 (18%) | 0,334 |
| Hyperlipidaemia, *n* (%) | 47 (94%) | 49 (98%) | 0,617 |
| BMI≥25kg/m<sup>2</sup>, *n* (%) | 35 (70%) | 39 (76%) | 0,713 |
| Previous MI, *n* (%) | 17 (34%) | 21 (42%) | 0,41 |
| Smoking status |  |  |  |
| Current smoker | 18 (36%) | 13 (26%) |  |
| Former smoker | 27 (54%) | 24 (48%) | *P* *=0,528 |

Abbreviations: BMI,
body mass index; CABG, coronary artery bypass grafting; iFR, instantaneous wave-free ratio; MI, myocardial infarction; n, number of patients; P, P value (Student t test); P\*, P value (chi-square $\chi^2$ test); SD, standard deviation.

### Instantaneous Wave-Free Ratio and Surgical Data

A total of 200 iFR measurements were conducted in cases of intermediate (50%–75%) stenosis (Table 3). During CABG procedures, surgery duration was comparable between the iFR-guided and angiography-guided groups (56.74 vs. 55.1 minutes; *P* = 0.62). Similarly, the duration of postoperative hospital stay did not differ significantly (11.58 vs. 10.95 days; *P* = 0.44). Notably, the iFR-guided group had a significantly lower number of grafts (median 3 [IQR: 2–3.25]) compared with the angiography-guided group (median 3 [IQR: 3–4]; *P* = 0.001).

**Table 3.**
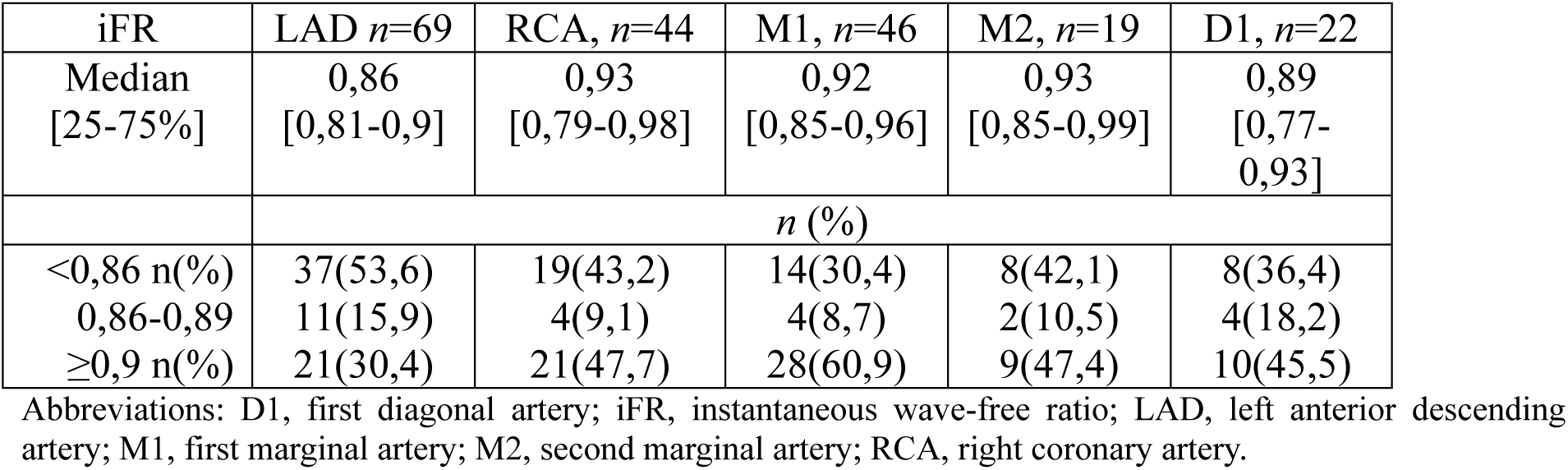
Instantaneous Wave-Free Ratio Values According to Coronary Artery.

| iFR | LAD *n*=69 | RCA, *n*=44 | M1, *n*=46 | M2, *n*=19 | D1, *n*=22 |
| --- | --- | --- | --- | --- | --- |
| Median<br>[25-75%] | 0,86<br>[0,81-0,9] | 0,93<br>[0,79-0,98] | 0,92<br>[0,85-0,96] | 0,93<br>[0,85-0,99] | 0,89<br>[0,77-0,93] |
|  | *n* (%) |  |  |  |  |
| <0,86 *n*(%) | 37(53,6) | 19(43,2) | 14(30,4) | 8(42,1) | 8(36,4) |
| 0,86-0,89 | 11(15,9) | 4(9,1) | 4(8,7) | 2(10,5) | 4(18,2) |
| ≥0,9 *n*(%) | 21(30,4) | 21(47,7) | 28(60,9) | 9(47,4) | 10(45,5) |

Abbreviations: D1, first diagonal artery; iFR, instantaneous wave-free ratio; LAD, left anterior descending artery; M1, first marginal artery; M2, second marginal artery; RCA, right coronary artery.

### Graft Patency During Follow-Up

Follow-up coronary computed tomography angiography (CCTA) was performed in 92 patients (92%) at 12 months and 78 patients (78%) at 36 months, evaluating a total of 253 grafts. At the 36-month follow-up, the rate of hypoperfusion or occlusion was significantly lower in the iFR-guided group compared with the angiography-guided group (15.15% [n = 20/132] vs. 32.82% [n = 43/131]; *P* = 0.007).

### LIMA-to-LAD Graft Patency

At the early follow-up (2 months), CCTA was performed for 94% of left internal mammary artery (LIMA) grafts where measurement by iFR in LAD was performed pre-CABG (for 64 out of 69pts.). Among these, 50 patent LIMAs had a median pre-CABG iFR of 0.855 (IQR: 0.785–0.892). Hypoperfusion was identified in 8 LIMAs (median iFR: 0.88 [IQR: 0.842–0.90]), while 7 LIMAs were occluded (median iFR: 0.91 [IQR: 0.88–0.96]; *P* = 0.004) (Figure 3).

**Figure 1.**
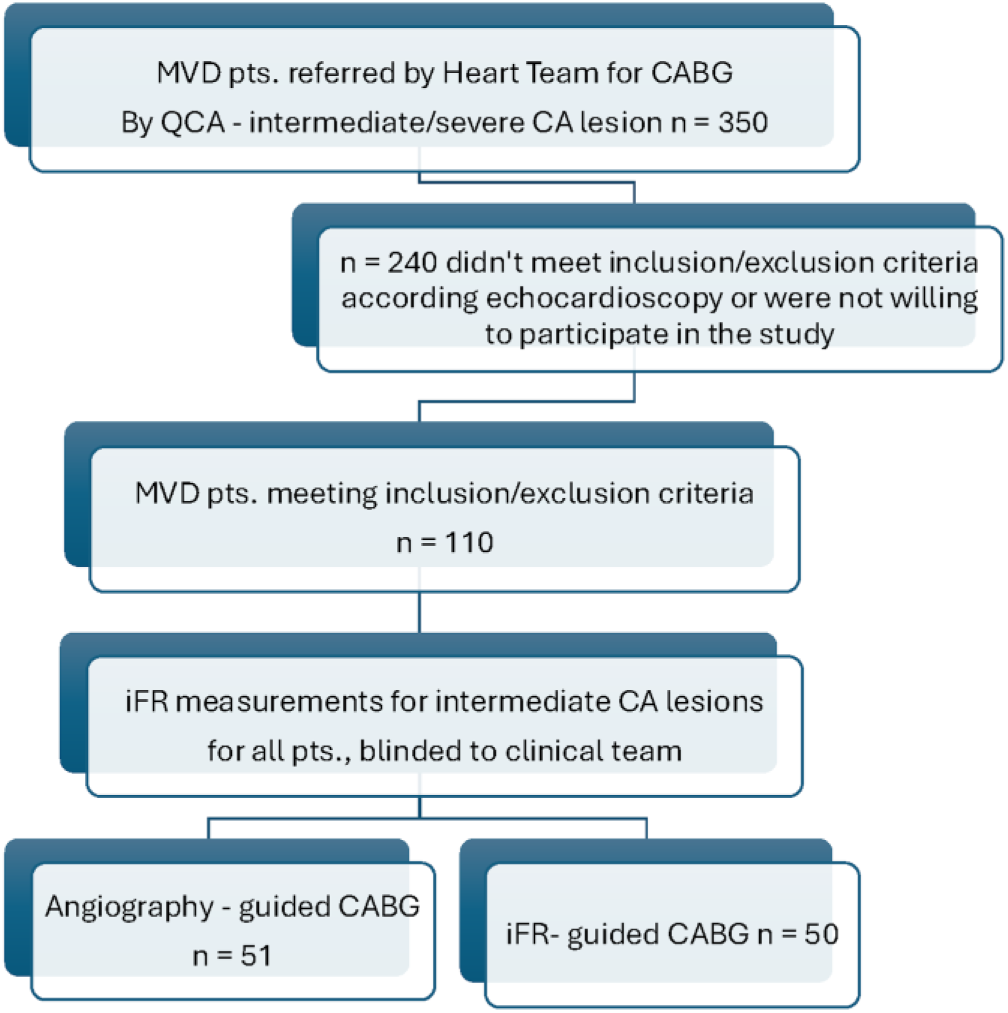
Study flowchart of patient enrolment, randomization, and treatment allocation. The flowchart illustrates the progression of participants through the iCABG study, including initial screening of patients with multivessel CAD, exclusion criteria application, and 1:1 randomization to either angiography-guided or iFR-guided CABG strategies. *n* indicates the number of patients in each cohort. Abbreviations: CABG, coronary artery bypass grafting; CAD, coronary artery disease; iFR, instantaneous wave-free ratio; MVD, multivessel disease; n, number of patients; QCA, quantitative coronary angiography.

**Figure 2.**
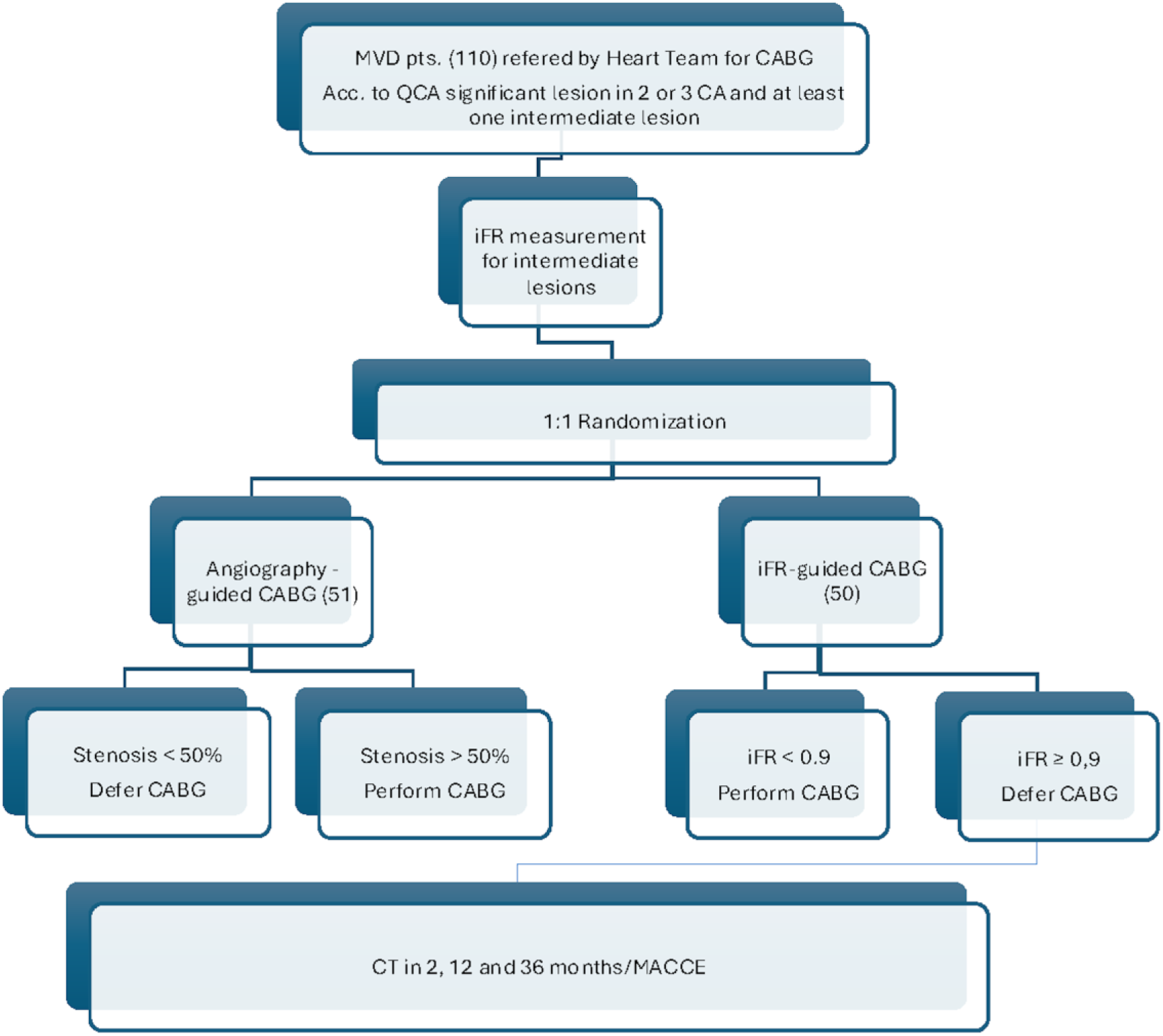
Study design and revascularization strategy flowchart. The flowchart illustrates the 1:1 randomization and treatment allocation for patients undergoing either angiography-guided or instantaneous wave-free ratio (iFR)-guided coronary artery bypass grafting (CABG). Scheduled clinical and imaging follow-up assessments were conducted at 2, 12, and 36 months postoperatively to evaluate graft patency and clinical outcomes. Abbreviations: CABG, coronary artery bypass grafting; CAD, coronary artery disease; CT, computed tomography; iFR, instantaneous wave-free ratio; MACE, major adverse cardiovascular events; MVD, multivessel disease; n, number of patients; QCA, quantitative coronary angiography.

**Figure 3.**
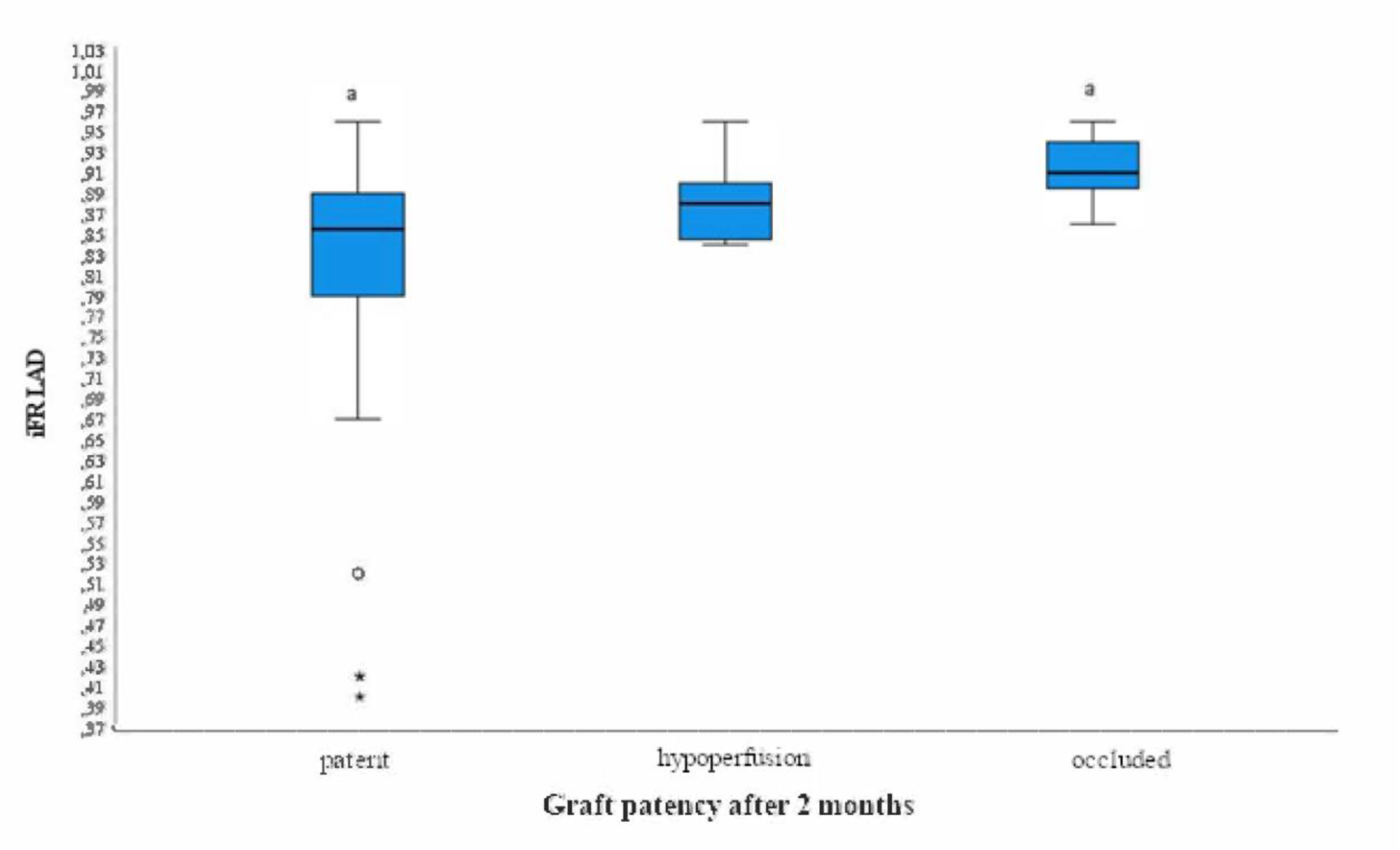
Preoperative Left Anterior Descending Artery (LAD) Instantaneous Wave-Free Ratio (iFR) Values Stratified by Graft Patency at 2-Month Follow-Up. Box plots represent the distribution of preoperative iFR values in the LAD artery based on graft patency status (patent, hypoperfused, or occluded) at 2 months post-CABG. Differences across groups were assessed using the Kruskal-Wallis test (*χ^2^*= 9.984; *df* = 2; *P* = 0.007). The observed difference in iFR values between groups was statistically significant (*P* = 0.004). Abbreviations: iFR, instantaneous wave-free ratio; LAD, left anterior descending artery.

At 12 months (90% follow-up), 32 LIMAs were patent (median iFR: 0.83 [IQR: 0.77–0.86]), 10 showed hypoperfusion (median iFR: 0.90 [IQR: 0.87–0.90]), and 14 were occluded (median iFR: 0.91 [IQR: 0.87–0.95]; *P* < 0.001) (Supplemental Figure S1).

At the 36-month follow-up (78% follow-up), LIMA-to-LAD graft patency was significantly higher in the iFR-guided group compared with the angiography-guided group (80.5% [n = 33/41] vs. 56.8% [n = 21/37]; *P* = 0.03; RR 1.42 [95% CI, 1.03–1.95]). Occluded or hypoperfused grafts were observed in 8 cases (19.5%) in the iFR-guided group versus 16 cases (43.2%) in the angiography-guided group (*P* < 0.001).

Subgroup analysis revealed that 90.0% of LIMA-to-LAD grafts were patent when pre-CABG iFR was <0.85, compared with 29.4% when iFR was >0.90 (*P* < 0.001). Graft patency was significantly higher for LAD lesions with iFR <0.85 compared to those with iFR 0.85 (90.0% vs. 41.9%; *P* < 0.005) (Figure 4). ROC curve analysis identified an optimal iFR threshold for predicting graft failure. At 12 months, a pre-CABG LAD iFR <0.85 was associated with a 3-fold higher likelihood of patency (OR 3.0 [95% CI, 1.89–4.76]). Conversely, at 36 months, the odds of LIMA-to-LAD graft failure (hypoperfusion/occlusion) were 12 times higher when the iFR exceeded 0.875 (OR 12.13 [95% CI, 3.14–46.95]) (Figures 5 and 6).

**Figure 4.**
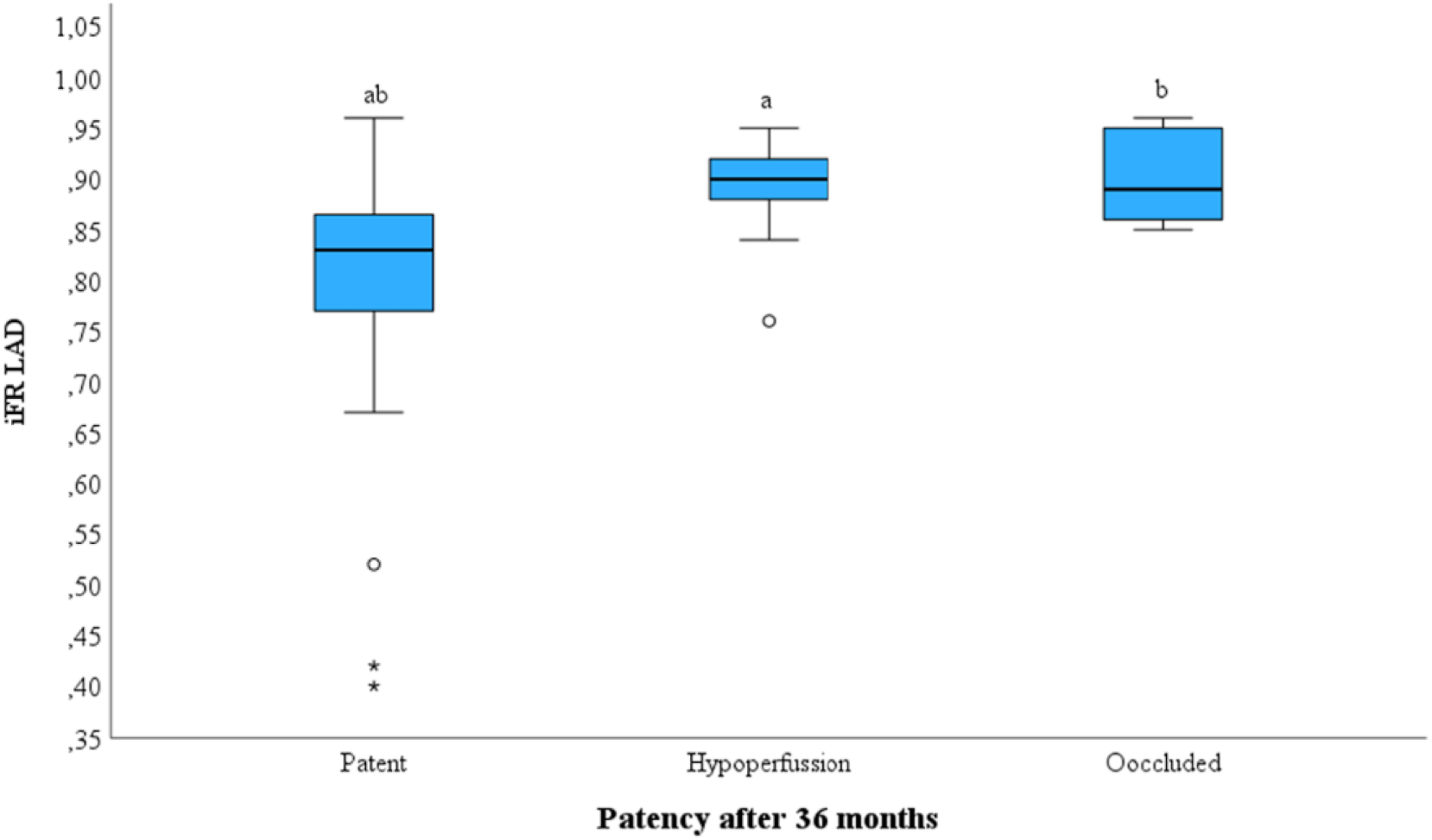
Preoperative Left Anterior Descending Artery (LAD) Instantaneous Wave-Free Ratio (iFR) Values and Left Internal Mammary Artery (LIMA) Graft Patency at 36-Month Follow-Up. Box plots illustrate the distribution of preoperative iFR values in the LAD artery, stratified by graft patency status (patent, hypoperfused, or occluded) at 36 months post-coronary artery bypass grafting. The plots depict the minimum, lower quartile, median, upper quartile, and maximum values, with outliers indicated individually. Differences across groups were assessed using the Kruskal-Wallis test (*χ^2^* = 13.758; *df* = 2; *P* < 0.001). Post-hoc pairwise comparisons demonstrated significant differences between the groups (all *P* < 0.05). Abbreviations: iFR, instantaneous wave-free ratio; LAD, left anterior descending artery; LIMA, left internal mammary artery.

**Figure 5.**
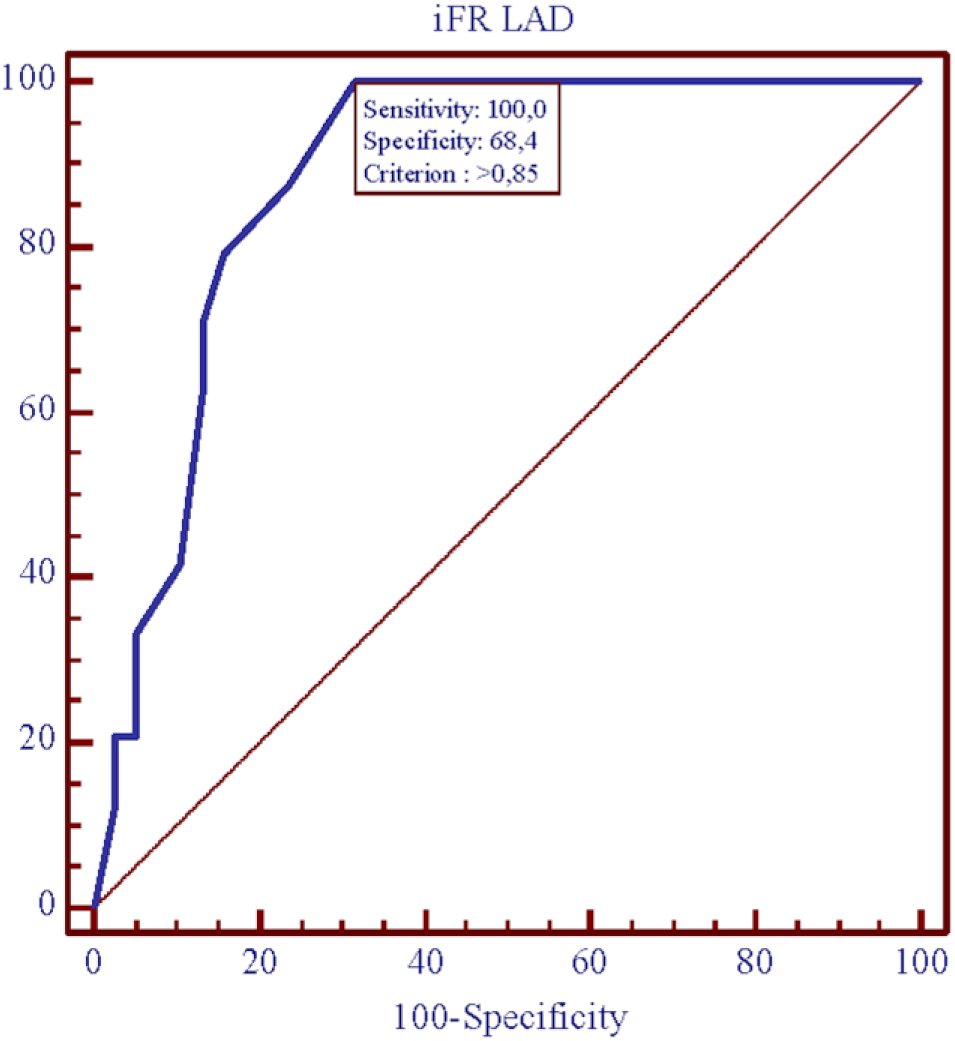
Receiver Operating Characteristic (ROC) Curve Analysis for Determining the Instantaneous Wave-Free Ratio (iFR) Threshold. The ROC curve illustrates the diagnostic performance of preoperative iFR in predicting graft failure (defined as hypoperfusion or occlusion) at the 12-month follow-up, as assessed by coronary computed tomography angiography. An iFR threshold of >0.85 was identified as the optimal cut-off for predicting compromised graft patency. Abbreviations: iFR, instantaneous wave-free ratio; LAD, left anterior descending artery; ROC, receiver operating characteristic.

**Figure 6.**
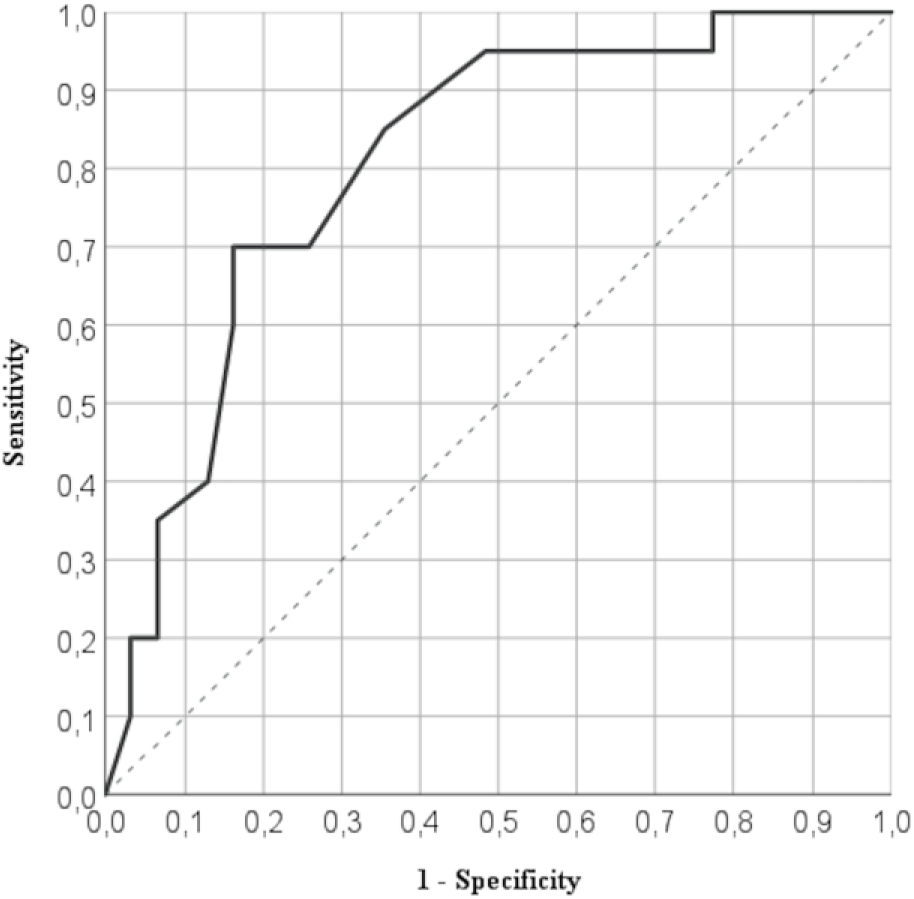
Receiver Operating Characteristic (ROC) Curve Analysis for Determining the Optimal Instantaneous Wave-Free Ratio (iFR) Threshold to Predict Graft Failure at 36 Months. The ROC curve illustrates the diagnostic performance of preoperative iFR in predicting long-term graft failure (defined as hypoperfusion or occlusion) at the 36-month follow-up, as assessed by coronary computed tomography angiography. This analysis confirms the sustained predictive value of the preoperative physiological assessment for graft patency over a 3-year period. Abbreviations: iFR, instantaneous wave-free ratio; ROC, receiver operating characteristic.

Patency of Saphenous Vein Grafts In a pooled analysis of saphenous vein grafts (diagonal, marginal, and right coronary arteries), the patency rate was significantly higher in the iFR-guided group (90.2% vs. 70.3%; *P* = 0.046) (Figure 7). The odds of a patent venous graft to the RCA at 12 months were 2.6 times higher when the iFR was >0.85 (OR 2.6 [95% CI, 1.307–5.171]) (Figure 8).

**Figure 7.**
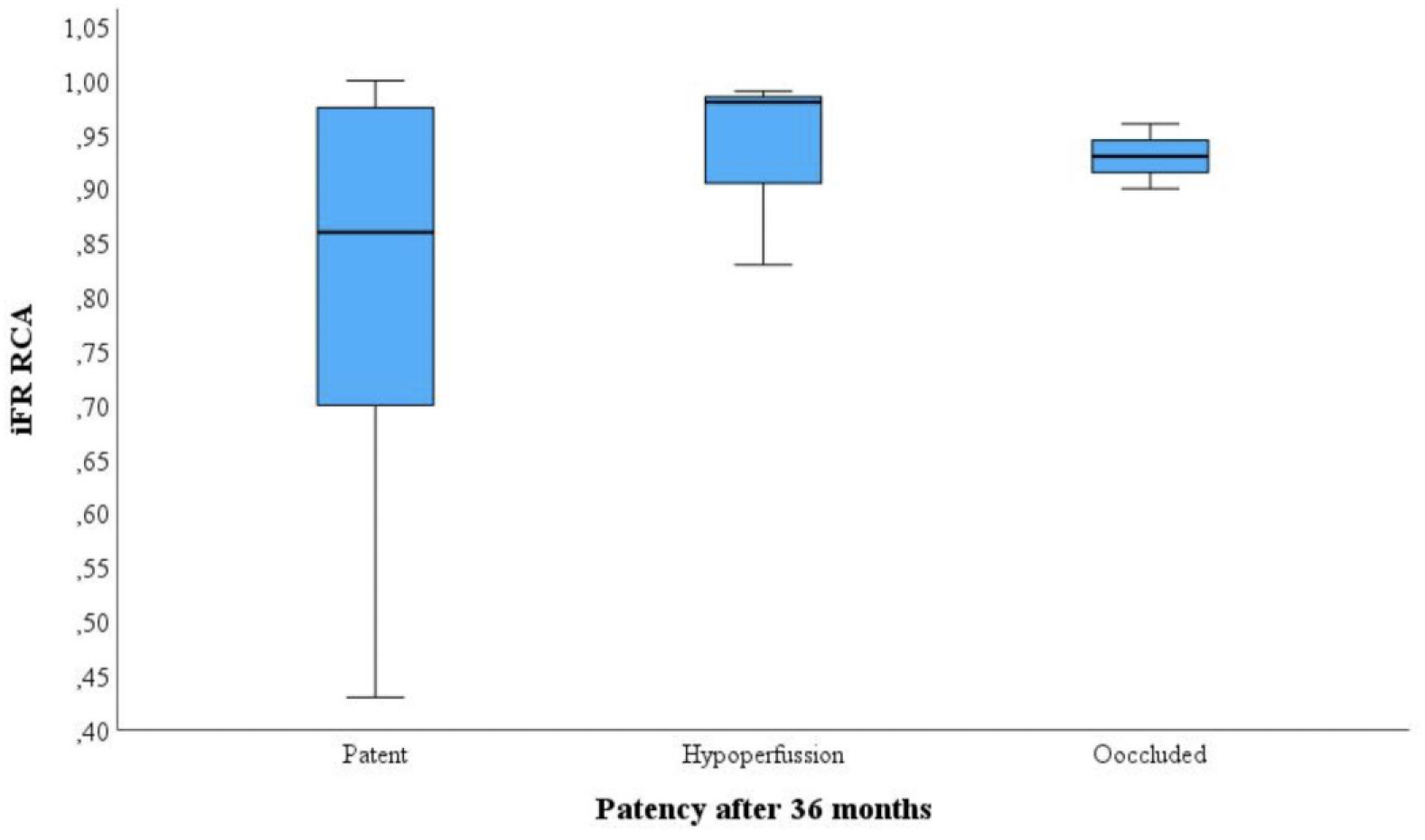
Preoperative Instantaneous Wave-Free Ratio (iFR) Values and Saphenous Vein Graft (SVG) Patency at 36-Month Follow-Up. Box plots illustrate the distribution of preoperative iFR values in coronary target vessels for SVG, categorized by patency status (patent, hypoperfused, or occluded) at 36 months post-coronary artery bypass grafting. Differences across groups were assessed using the Kruskal-Wallis test (*χ^2^* = 1.525; *df* = 2; *P* = 0.466), indicating no statistically significant association between preoperative iFR and long-term vein graft patency. Abbreviations: CABG, coronary artery bypass grafting; iFR, instantaneous wave-free ratio; SVG, saphenous vein graft.

**Figure 8.**
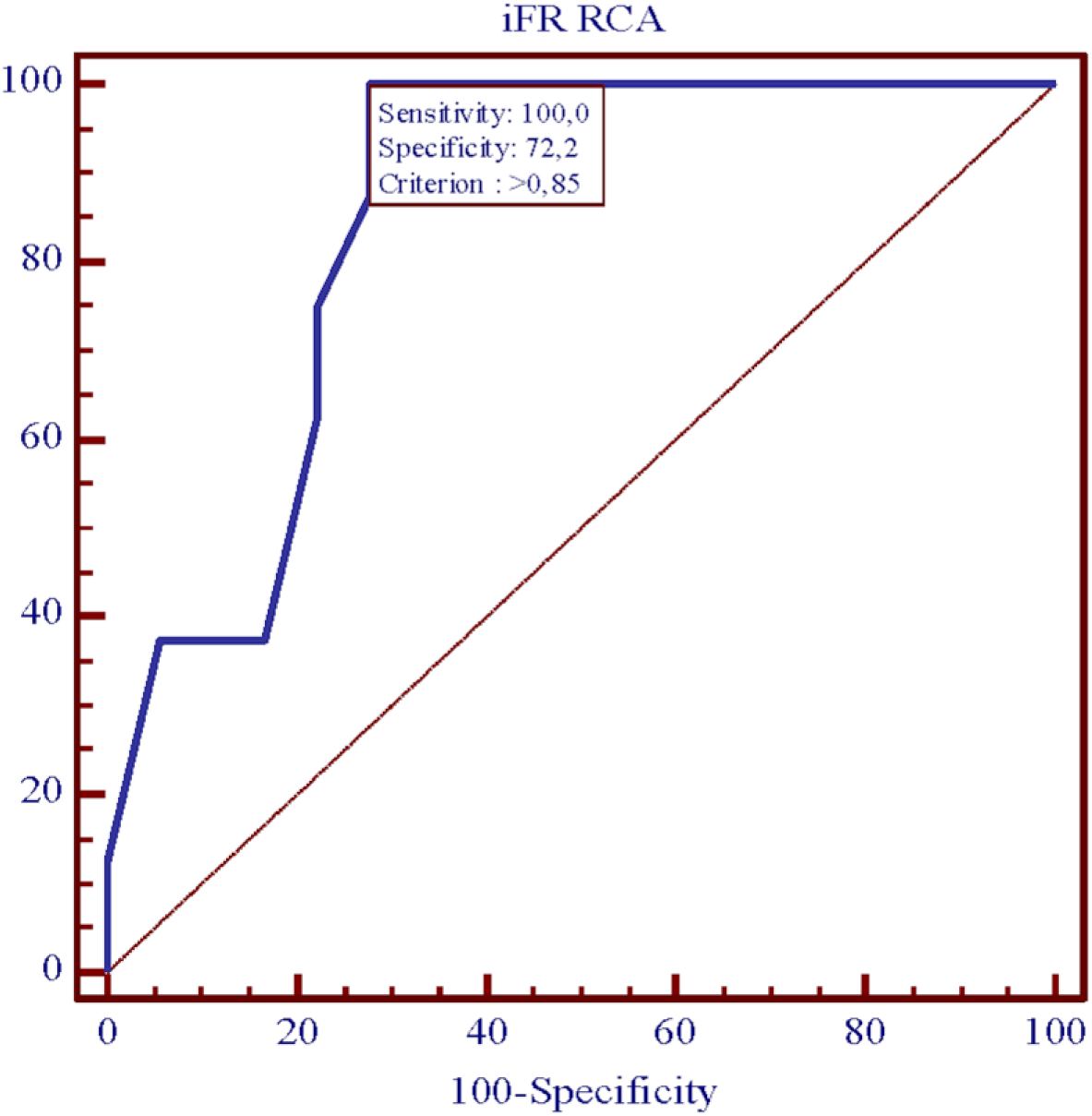
Receiver Operating Characteristic (ROC) Curve Analysis for Determining the Optimal Instantaneous Wave-Free Ratio (iFR) Threshold to Predict Right Coronary Artery (RCA) Graft Patency. The ROC curve illustrates the diagnostic performance of preoperative iFR in predicting RCA graft patency. An iFR threshold of was identified as the optimal cut-off value, demonstrating a sensitivity of 100.0% and a specificity of 72.2% for predicting graft hypoperfusion or occlusion. These findings align with the physiological impact of competitive flow on RCA graft longevity. Abbreviations: iFR, instantaneous wave-free ratio; RCA, right coronary artery; ROC, receiver operating characteristic.

### Clinical Outcomes

No significant difference in MACCE rates was observed between groups at 36 months (14 events in the iFR group vs. 10 events in the angiography group; *P* = 0.48, corresponding to 28% vs. 20% respectively). Similarly, anginal status (CCS Class 1-2) did not differ significantly (*P* = 0.57).

Overall mortality was attributed to non-cardiovascular causes: malignancy (n = 2), COVID-19 (n = 1), and accidents (n = 1). Three cerebrovascular strokes were recorded (two early post-CABG, one at 6 months). No myocardial infarctions were reported. A single case of target vessel revascularization (TVR) occurred: a patient with a pre-CABG LAD iFR of 0.76 developed LIMA occlusion at 2 months (repeat iFR 0.75, subsequently undergoing successful PCI of the LAD.

## DISCUSSION

This study aimed to compare the outcomes of iFR-guided versus angiography-guided CABG by assessing graft patency, MACCE, and clinical outcomes at a 36-month follow-up. Our findings demonstrate that physiological guidance using iFR significantly improves graft patency following CABG compared with angiography-guided revascularization alone. This benefit was observed for both arterial and venous grafts and persisted at mid-term follow-up. While iFR-guided CABG resulted in significantly higher graft patency rates, especially for intermediate-grade stenosis, there was no significant difference in the occurrence of MACCE between the two groups.

The benefit of iFR has been demonstrated in several multicentre prospective studies, which found that utilizing iFR-based assessment protocols improves both early and late outcomes following percutaneous coronary intervention (PCI) [12, 13]. The randomized SWEDEHEART and DEFINE-FLAIR trials, evaluating 1-year and 5-year outcomes, demonstrated that iFR is non-inferior to fractional flow reserve (FFR) for determining the extent of revascularization [14, 15]. Our study provides prospective randomized evidence supporting the extension of these physiological lesion assessment principles into surgical practice, reinforcing the concept that coronary physiology should guide all modes of revascularization.

### Clinical and Procedural Outcomes

Several studies have demonstrated that FFR-guided surgical revascularization is associated with a lower number of grafts, shorter aortic clamp time, and reduced inpatient stay; however, no difference was found in 12-month MACCE rates compared to angiography-guided CABG [16]. In our study, the in-hospital stay duration did not differ significantly between the iFR-guided and angiography-guided groups (11.58 vs. 10.95 days; *P* = 0.44), aligning with Bruno et al.’s meta-analysis [17]. However, we observed a significantly lower number of grafts in the iFR-guided group (mean 3.0) compared to the angiography-guided group (mean 3.5; *P* = 0.001), consistent with findings in FFR-guided procedures [17].

Regarding MACCE, our study found no statistically significant difference between the groups (*P* = 0.48), mirroring the results of the GRAFFITI trial [18]. In contrast, Fournier et al. reported that FFR-guided CABG was associated with a lower incidence of MACCE [19], although their data were derived from a retrospective analysis. While our 36-month data did not show a MACCE benefit, the superior long-term graft patency observed with iFR guidance may translate into improved clinical outcomes over an extended follow-up period.

### Mechanistic Insights and Prediction of Graft Patency

A primary objective of this study was to evaluate the utility of preoperative iFR in predicting postoperative graft patency. Graft failure following CABG is multifactorial, with competitive native coronary flow representing as a key determinant. We observed that grafts were frequently occluded or hypoperfused when the preoperative iFR value was physiologically non-significant, including the “grey zone” (iFR >0.85). In the presence of non-significant stenosis, concurrent flow between the antegrade native circulation and the graft is the primary driver of early conduit failure [22]. This is supported by the PREVENT IV trial, where LAD stenosis <75% was identified as a risk factor for LIMA-LAD graft failure [23]. Other FFR-based studies have similarly shown that LIMA-LAD patency is higher when the target lesion is hemodynamically significant [24].

Notably, the correlation between physiological significance and graft patency was not confirmed in the GRAFFITI trial [18]. However, that trial had specific limitations, including the inclusion of “grey zone” FFR values (0.75–0.80) and patients with acute coronary syndromes, which may have confounded the patency distribution. Our findings, utilizing the ROC test, determined that an iFR >0.85 is a potent predictor of failure, exhibiting 100% sensitivity and 68.4% specificity for LIMA-LAD grafts, and 100% sensitivity and 72.2% specificity for all saphenous vein grafts at 36 months. These results support the hypothesis that physiology-guided target selection optimizes conduit durability by avoiding non-flow-limiting stenoses where competitive flow would otherwise promote thrombosis.

### Clinical Implications and Study Limitations

Improved graft patency has profound long-term implications, as conduit failure is traditionally associated with recurrent angina and the need for repeat revascularization. Physiology-guided CABG may therefore reduce the healthcare burden associated with late graft failure. This represents an important shift in surgical planning, providing a more objective basis for target selection.

Several limitations should be acknowledged:

1. This was a single-center study; however, standardized surgical and imaging protocols ensured data consistency.
2. The sample size was moderate, which may limit the statistical power to detect differences in clinical endpoints such as MACCE.
3. Follow-up was limited to 36 months. Longer-term surveillance is required to determine if the patency benefits eventually manifest as superior clinical outcomes.

## CONCLUSIONS

iFR-guided CABG significantly improves graft patency compared with angiography-guided CABG. Physiological assessment under the resting conditions enables the identification of hemodynamically significant lesions and allows avoiding unnecessary grafting, representing a significant advancement in the optimization of coronary artery bypass surgery outcomes or CABG deferring at all.

## Data Availability

The data that support the findings of this study are available from the corresponding author, Rasa Ordiene, upon reasonable request.

## ABBREVATONS

CABG: coronary artery bypass grafting
CCTA: coronary computed tomography angiography
FFR: fractional flow reserve
iFR: instantaneous wave-free ratio
LAD: left anterior descending artery
LIMA: left internal mammary artery
MACCE: major adverse cardiac and cerebrovascular events

## REFERENCES

1. Sen S, Escaned J, Malik IS, et al. Development and validation of a new adenosine-independent index of stenosis severity from coronary wave-intensity analysis: results of the ADVISE study. J Am Coll Cardiol. 2012;59:1392–1402.

2. Lee JM, Choi KH, Koo BK, et al. Comparison of major adverse cardiac events between instantaneous wave-free ratio and fractional flow reserve-guided strategy. JAMA Cardiol. 2019;4:857–864.

3. Gotberg M, Christiansen EH, Gudmundsdottir IJ, et al. Instantaneous wave-free ratio versus fractional flow reserve to guide PCI. N Engl J Med. 2017;376:1813–1823.

4. Modi BN, Rahman H, Kaier T, et al. Revisiting optimal fractional flow reserve and instantaneous wave-free ratio thresholds. Circ Cardiovasc Interv. 2018;11:e007041.

5. Botman CJ, Schonberger J, Koolen S, et al. Does stenosis severity influence bypass graft patency? Ann Thorac Surg. 2007;83:2093–2097.

6. Tolegenuly A, Ordiene R, Mamedov A, Unikas R, Benetis R. Correlation between iFR and intraoperative graft flow measurement. Medicina (Kaunas). 2020;56:714.

7. Sabik JF, Blackstone EH. Coronary artery bypass graft patency and competitive flow. J Am Coll Cardiol. 2008;51:126–128.

8. Neumann FJ, Sousa-Uva M, Ahlsson A, et al. ESC/EACTS guidelines on myocardial revascularization. Eur Heart J. 2019;40:87–165.

9. Gaudino M, Antoniades C, Benedetto U, et al. Mechanisms, consequences, and prevention of coronary graft failure. Circulation. 2017;136:1749–1764.

10. van de Hoef TP, Meuwissen M, Escaned J, et al. Head-to-head comparison of iFR and FFR. EuroIntervention. 2015;11:914–925.

11. Balacumaraswami L, Taggart DP. Intraoperative imaging techniques to assess graft patency. Ann Thorac Surg. 2007;83:2251–2257.

12. Otsuka F, Yahagi K, Sakakura K, Virmani R. Why is the mammary artery so special? J Am Heart Assoc. 2013;2:e000099.

13. Toth GG, De Bruyne B, Kala P, et al. Graft patency after FFR-guided vs angiography-guided CABG. EuroIntervention. 2019;15:e999–e1005.

14. Harskamp RE, Alexander JH, Ferguson TB, et al. Frequency and predictors of internal mammary artery graft failure. Circulation. 2016;133:131–138.

15. Escaned J, Ryan N, Mejia-Renteria H, et al. Safety of the deferral of coronary revascularization based on iFR and FFR. JACC Cardiovasc Interv. 2018;11:1437–1449.

16. Bruno F, D’Ascenzo F, Marengo G, et al. FFR-guided vs angiography-guided CABG: systematic review. Eur Heart J. 2020;41(Suppl 2):ehaa946.3601.

17. Fournier S, Toth GG, Colaiori I, et al. Long-term patency after FFR-guided CABG. Circ Cardiovasc Interv. 2019;12:e007712.

18. Kang DY, Ahn JM, Lee CH, et al. Deferred vs performed revascularization based on FFR. Eur Heart J. 2018;39:1610–1619.

19. Berger A, MacCarthy PA, Siebert U, et al. Long-term patency of internal mammary artery grafts. Circulation. 2004;110:II36–II40.

20. Fournier S, Toth GG, De Bruyne B, et al. Six-year follow-up of FFR-guided CABG. Circ Cardiovasc Interv. 2018;11:e006368.

21. He GW, Fan L, Grove KL, et al. Endothelial nitric oxide synthase expression in arterial grafts. Ann Thorac Surg. 2011;92:845–850.

22. Davies JE, Sen S, Dehbi HM, et al. Use of instantaneous wave-free ratio or fractional flow reserve in PCI. N Engl J Med. 2017;376:1824–1834.

